# COVID-19 and Environmental factors. A PRISMA-compliant systematic review

**DOI:** 10.1101/2020.05.10.20069732

**Authors:** I. Chatziprodromidou, T. Apostolou, A. Vantarakis

## Abstract

The emergence of a novel human coronavirus, SARS-CoV-2, has become a global health concern causing severe respiratory tract infections in humans. Human-to-human transmissions have been described with incubation times between 2-10 days, facilitating its airborne spread via droplets. The impact of environmental factors on the coronavirus disease 2019 (COVID-19) outbreak is under consideration. We therefore reviewed the literature on all available information about the impact of environmental factors on human coronaviruses. Temperature, humidity and other environmental factors have been recorded as environmental drivers of the COVID-19 outbreak in China and in other countries. Higher temperatures might be positive to decrease the COVID-19 incidence. In our review, the analysis of 23 studies show evidence that high temperature and high humidity reduce the COVID-19 transmission. However, further studies concerning other environmental (namely meteorological) factors’ role should be conducted in order to further prove this correlation.

As no specific therapies are available for SARS-CoV-2, early containment and prevention of further spread will be crucial to stop the ongoing outbreak and to control this novel infectious thread.

## Introduction

A novel coronavirus (SARS-CoV-2) has recently emerged from China with a total of 45171 confirmed cases of pneumonia (as of February 12, 2020). Coronaviruses (CoVs) most commonly cause mild illness; but have occasionally, in recent years, led to major outbreaks of human disease. Approximately ten years after SARS, another novel, highly pathogenic CoV, in December 2019, SARS-CoV-2, a novel CoV, was identified in the City of Wuhan, Hubei Province, a major transport hub of central China. The earliest COVID-19 cases were linked to a large seafood market in Wuhan, initially suggesting a direct food source transmission pathway (Thompson 2020). Along with Severe Acute Respiratory Syndrome (SARS) coronavirus and Middle East Respiratory Syndrome (MERS) coronavirus (de Wit et al. 2016) (Peeri et al. 2020), this is the third highly pathogenic human coronavirus that has emerged in the last two decades. In the months since the identification of the initial cases, COVID-19 has spread to 180 countries and territories and there are approximately 664,564 confirmed cases and 30,890 deaths (as of 29 March 2020).

Person-to-person transmission was confirmed as one of the main mechanisms of COVID-19 spread (Chan et al. 2020). The modes of transmission have been identified as host-to-human and human-to-human. Increased spread of SARS-CoV-2 causing COVID-19 infections worldwide has brought increased attention and fears surrounding the prevention and control of SAR-CoV-2 from both the scientific community and the general public. While many of the precautions typical for halting the spread of respiratory viruses are being implemented, other less understood transmission pathways should also be considered and addressed to reduce further spread.

The role of environment and its mediated pathways for infection by other pathogens have been a concern for decades. Substantial research into the presence, abundance, diversity, function, survival and transmission of microorganisms in the environment has taken place in recent years. There is preliminary evidence that environmentally mediated transmission may be possible; additionally, that COVID-2 could be affected by environmental factors such as seasonality, temperature, humidity (Tan et al. 2005) (Shaman and Kohn 2009) (Geller, Varbanov, and Duval 2012).

The aim of the review was, therefore, to summarize all available data on the impact of environmental factors on the survival of all coronaviruses including emerging SARS-CoV and MERS CoV.

## Methodology

Methodology of this systematic review and inclusion criteria were indicated in advance and recorded in *a priori* protocol in order to determine the rationale, the objectives, the eligibility and selection criteria, the search strategy and the study selection process of this systematic review. However, due to emergency of the subject and due to the pandemic awareness for COVID-19, this systematic review was not registered with PROSPERO (International Prospective Register of Systematic Reviews).

### Eligibility criteria

All study design types were considered in this systematic review. Reviews were not included but screened for any information within the scope of this review. No language, publication status or publication year restrictions were imposed. As because of the COVID-19 emergency state, even not proofread publications were included in our study. All non-English studies, including Chinese, Japanese and French were translated via Google translator and were included in this systematic review. Although COVID-19 concerns years 2019 and 2020, no year of publication limit was applied, in order to exploit valuable information concerning the coronavirus relationship with environmental factors, as indicated by the past SARS and MERS lessons. All studies included, concern human coronavirus strains of various types. This systematic review was limited to studies focusing to environmental factors’ impact on COVID-19. Searched experts’ and researchers’ opinions were not handed in this study.

The selection criteria developed a priori are described below:

- Year of publication
- Country of epidemics
- Continent of epidemics
- Environmental factor
- Assessing method

### Information sources

The search strategy and analysis process were conducted according to the Preferred Reporting Items for Systematic Reviews and Meta-analyses (PRISMA) statement for systematic reviews (Liberati et al. 2009; Page et al. 2018). Titles and abstracts of the retrieved articles were screened, while full length articles were evaluated for eligibility and were further acquired via SwetsWise Online Content. Search for articles was applied in three electronic databases: Google Scholar, PubMed & Springerlink. Google Scholar was our starting point. No unpublished information obtained. The literature search was performed from 25 to 28 of March 2020.

### Search

The following terms were used to search all databases, always in combination with “coronavirus” and “COVID-19”: “environmental factors”, “clima”, “temperature”, “humidity”, “absolute humidity”, “relative humidity”, “wind speed”, “wind power”, “precipitation”, “rainfall”. The search strategy was conducted by IPC and was peer-reviewed by AV as part of the systematic review process.

### Study selection

Eligibility assessment procedure was performed in standardized and independent manner, primary by two authors (IPC and AV), to analyze and validate all relevant data to the top under discussion. Disagreements were resolved through discussion among all authors and resulted in a final consensus. After excluding records upon the eligibility criteria set, we screened all titles and abstracts of the retrieved studies, although full text review proves also to be necessary for further consideration.

### Data Collection process

A data extraction sheet was developed in order to summarize the evidence of this systematic review, based on the Cochrane Consumers and Communication Review Group’ data extraction template for included studies (Cochrane Consumers and Communication 2016). This was pilot tested on the first ten randomly selected studies and no refinement was needed. One author (IPC) extracted all proper data from the included studies and another author (AV) checked all the extracted data. No disagreements arose. In order to ascertain duplicate publications, we used the tool “check for duplicates” of Mendeley Desktop software (Version 1.19.4).

### Assessment of study quality

To ascertain the validity of the included studies, two reviewers (IPC and AV) in a blind manner and independently scored the included papers’ quality upon the Strengthening the Reporting of Observational Studies in Epidemiology (STROBE) checklist (Von Elm et al. 2014). Both reviewers independently scored each included paper’s quality and all studies received a score that ranged from 0 to 22 points by each reviewer. Base up on a criterion included in the initial protocol of the study, the scores between the reviewers should not differ from one to another reviewer by more than 2 points. In order to generate a final score, both scores of the reviewers were averaged.

### Planned methods of analysis

In order to handle data and combine the results of all the included studies, we used SPSS (Statistical Package for the Social Sciences) (IBM SPSS Statistics for Macintosh, Version 25.0 n.d.) or R software (R Development Core Team 2013).

## Results

### Study selection and characteristics

The search of Google Scholar, Springerlink and PubMed provided a total of 14640, 51 and 28 articles, respectively. From the initially obtained 14719 articles, 8499 were excluded as duplicated ones upon “Check for duplicates” tool of Mendeley Desktop. The remaining 6220 articles were assessed for eligibility and a total of 6007 articles were discarded because based on a detailed evaluation of abstracts, they did not meet the eligibility criteria set and concerned: a) 2457 discussed clinical and epidemiological considerations of COVID-19, b) 813 discussed environmental factors associated with other diseases, c) 760 discussed ethical considerations od COVID-19, d) 743 discussed transmission dynamics of COVID-19, e) 655 discussed diagnostic and management outbreak investigations of COVID-19, f) 500 discussed prospects and advances in designing and developing vaccine and immuno-therapeutics of COVID-19 and g) 79 discussed socio-economic impact of COVID-19. Of the remaining 213 articles, 124 were excluded because they did not meet the inclusion criteria. The full text of the rest 89 articles was evaluated in further detail. Finally, 23 were included for further analysis in this systematic review (Figure 1).

**Figure 1.**
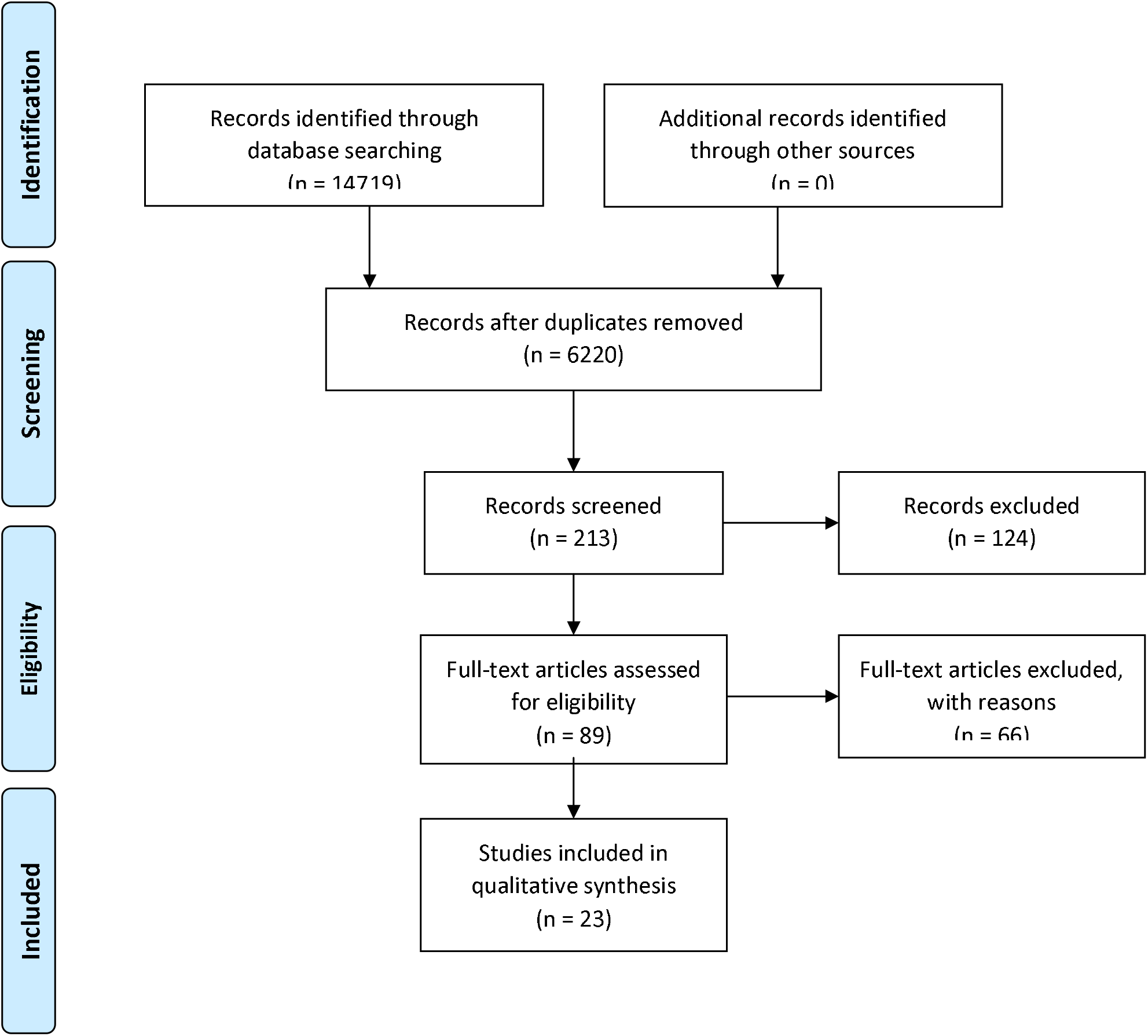
Flow diagram of the study selection process

All 23 studies selected for this systematic review were published in 2020 and in English. 65.2% of the included studies reported China, 26.1% did not mention a certain country of epidemics, 4.3% concerned the epicentre of the disease, namely Iran, Italy, South Korea, etc. and 4.3% concerned Singapore. Upon continent of epidemics, Asia hold the leads with 69,6%, followed by Africa with 4.3%, whereas almost 21.7% did not mention specific continent and 4.3% refers to mixed continents (Asia, Europe, etc.). All included studies assessed the role of various environmental factors on transmission rates of the COVID-19.

In 24.1% of the studies, temperature was assessed for its impact on COVID-19, followed by humidity (11.1%), absolute humidity (5.6%), rainfall/precipitation (5.6%), relative humidity (5.6%), travel (5.6%), air travel (3.7%), wind speed/power (3.7%), latitude (3.7%), built environment (1.9%), general lockdown (1.9%), visibility (1.9%), specific humidity (1.9%), airborne dust (1.9%), air pollution (1.9%), chemical pollution (1.9%), air index (1.9%), atmospheric radiation (1.9%), cloud cover (1.9%), precipitation of the driest month (1.9%), mean temperature of the wettest quarter (1.9%), isothermality (day-to-night temperatures difference relative to the summer-to-winter annual difference) (1.9%), annual mean temperature (1.9%), mean diurnal range (1.9%), minimum temperature of the coldest month (1.9%) and precipitation of the coldest quarter (1.9%).

In order to examine the association between those environmental factors and COVID-19, most of the studies employed the review method (20.4%), followed by maximum entropy model (13%), the model (11.1%), dynamical model and ERA-5 reanalysis (9.3%), the statistical modeling Loess regression (Generalized-linear or non-linear model) (7.4%), the R proxy method (5.6%), the R reproductive number (3.7%), the One-way ANOVA followed by a post-hoc Tukey’s HSD test (3.7%), the distributed lag log-linear model (3.7%), the linear regression model (3.7%), the global meta-population disease transmission model (3.7%), the Mann-Whitney U test (3.7%), the mathematical model (3.7%), the restricted cubic spline function and the generalized linear mixture model (3.7%) and the multivariate analysis (3.7%). Figure 2 displays the environmental factor assessed, combined with the assessing method and the country of epidemics it concerns. Detailed characteristics of the studies included, like author, title and year of publication, country and continent of the study, method of assessing the impact of the environmental factors and the outcome variable are described in Table 1.

**Figure 2.**
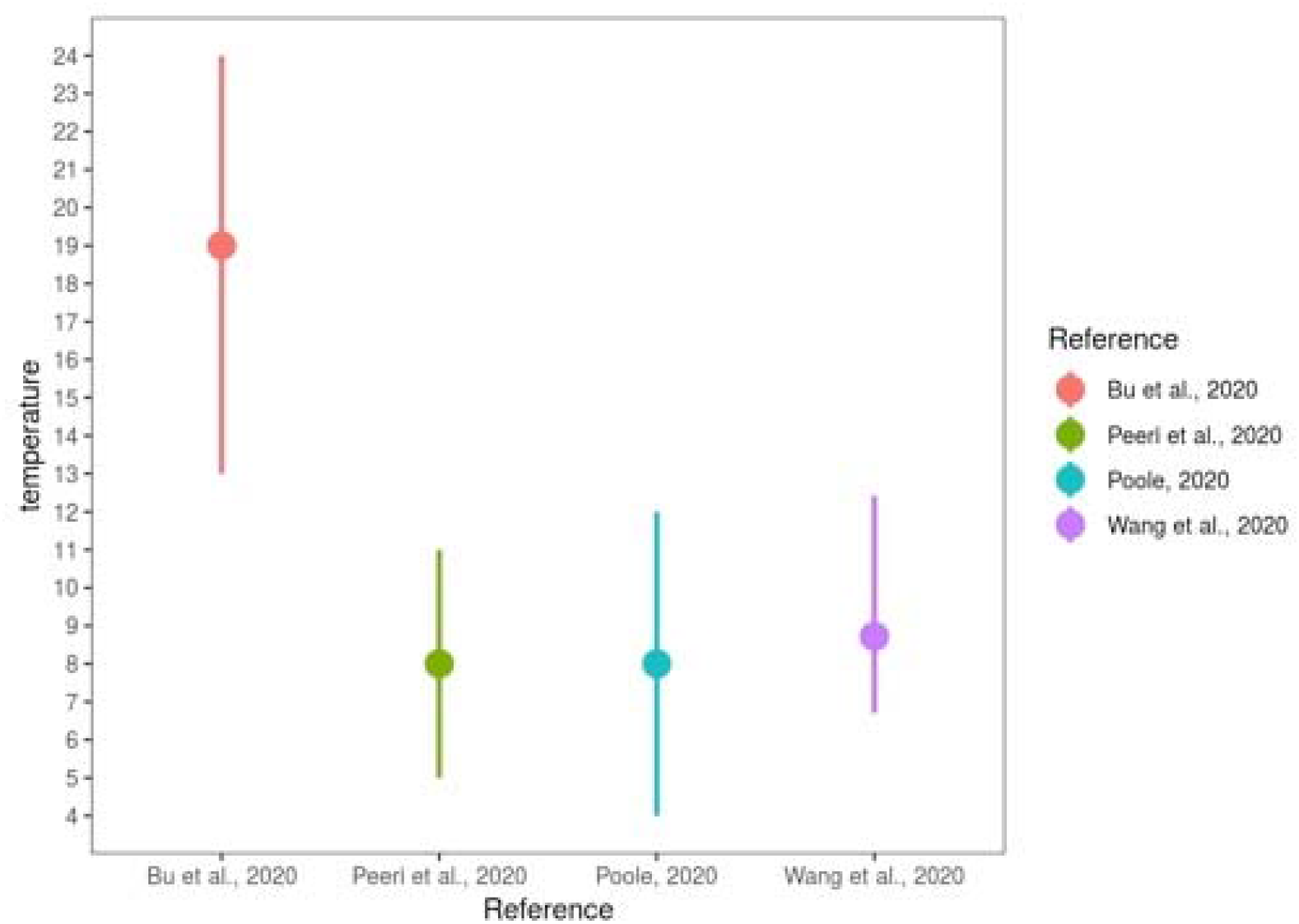
Environmental factors associated with the assessing methods the country of epidemics

**Table 1.** Characteristics of the studies included in this systematic review

|  | Author/year | Country of epidemics | Continent of epidemics | Assessing method | Environmental factor assessed |
| --- | --- | --- | --- | --- | --- |
| 1 | Gilbert et al., 2020 | Not mentioned | Africa | Multivariate analysis | Air travel |
| 2 | Wang et al., 2020 | China | Asia | Restricted cubic spline function & Generalized linear mixture model | Temperature |
| 3 | Luo et al., 2020 | China | Asia | Estimation of a proxy for the reproductive number | Absolute humidity |
| 4 | Bonilla-Aldana et al., 2020 | Not mentioned | Not mentioned | Review | Temperature<br>Rainfall/precipitation<br>Humidity |
| 5 | Poirier et al., 2020 | Not mentioned | Not mentioned | Estimation of a proxy for the reproductive number | Temperature<br>Humidity |
| 6 | Dietz et al., 2020 | Not mentioned | Not mentioned | Review | Built environment |
| 7 | Oliveiros et al., 2020 | China | Asia | Linear regression model | Temperature<br>Humidity |
| 8 | Shi et al., 2020 | China | Asia | 3 Distributed lag loglinear models | Temperature<br>Absolute humidity |
| 9 | Lau et al., 2020 | China | Asia | One-way ANOVA followed by a post-hoc Tukey's HSD test | Air travel<br>General lockdown |
| 10 | Chen et al., 2020 | China | Asia | Statistical modelling: Loess regression (Generalized-linear or non-linear model) | Temperature<br>Visibility<br>Wind speed/power<br>Relative humidity |
| 11 | Sun et al., 2020 | China | Asia | Review | Temperature<br>Humidity<br>Rainfall/Precipitation |
| 12 | Gostic et al., 2020 | Not mentioned | Not mentioned | Model | Travel |
| 13 | Peeri et al., 2020 | Epicentre (Iran, Italy, etc.) | Epicentre (Iran, Italy, etc.) | ERA-5 reanalysis | Temperature<br>Absolute humidity<br>Humidity<br>Specific humidity<br>Latitude |
| 14 | Qu et al., 2020 | Not mentioned | Not mentioned | Review | Airborne dust<br>Air pollution<br>Chemical pollution |
| 15 | Gupta, 2020 | China | Asia | Mathematical model | Temperature |
| 16 | Cai et al., | China | Asia | Mann-Whitney U test | Temperature |
| 17 | Chinazzi et al., 2020 | China | Asia | Global metapopulation disease transmission model | Travel |
| 18 | Lee et al., 2020 | Singapore | Asia | Review | Travel |
| 19 | Wang et al., 2020 | China | Asia | Estimation of a proxy for the reproductive number | Temperature<br>Relative humidity |
| 20 | Jiwei et al., 2020 | China | Asia | Dynamical model | Temperature<br>Wind speed/power<br>Rainfall/precipitation<br>Relative humidity<br>Air index |
| 21 | Poole, 2020 | Worldwide | Worldwide | Model | Temperature<br>Atmospheric pollution<br>Humidity<br>Cloud cover<br>Latitude |
| 22 | Bariotakis et al., 2020 | Worldwide | Worldwide | Maximum entropy model | Precipitation<br>Isothermality (day-to-night temperatures difference relative to the summer-to-winter (annual) difference)<br>Min temperature of the coldest month<br>Mean diurnal range<br>Mean temperature of wettest quarter<br>Annual mean temperature |
| 23 | Bu et al., 2020 | China | Asia | Review | Temperature<br>Humidity<br>Rainfall/Precipitation |

Figure 2 displays the exact temperature range proposed by certain studies included in this systematic review, in which virus survival is facilitated.

### Results of individual studies

Gilbert et al. used the volume of air travel concerning the flights from the infected China provinces (Guangdong, Fujan and the city Beijing) to Africa and concluded that there are 2 identified clusters of African countries: a) those that have the higher importation risk of exposure to COVID-19, which have moderate to high capacity to face an outbreak, like Egypt, Algeria and South Africa and b) those that are at moderate risk and have high vulnerability and variable capacity (Gilbert et al. 2020). Mao et al. concluded that temperature has a non-linear dose response relationship with COVID-19 transmission, whereas there is a specific temperature range, in which virus transmission is facilitated and this might also explain the emergence of the epidemic in Wuhan city. Wang et al. suggest that regions with lower temperature records should take even stricter measures in order to prevent future outbreaks (Wang et al. 2020). Luo et al. and Poirier et al. suggest that changes in weather conditions alone, namely increase of humidity and temperature (which are usually met in spring and summer seasons) may not suffice to decrease the number of cases, if no proper public health interventions are adopted (Luo et al. 2020) (Poirier et al. 2020). Bonilla-Aldana et al. propose that temperature, rainfall and humidity may play a significant role in the virus transmission, as occurs for many zoonotic diseases (Bonilla-Aldana, Dhama, and Rodriguez-Morales 2020). Dietz et al. combined current literature and built environment and assessed its role in COVID-19 transmission, suggesting that built environment plays a significant role in the disease control and mediation and this may be taken into consideration in the building design market(Conraths et al. 2015). Oliveiros et al. regressed the doubling time of COVID-19 cases with the aid of temperature and humidity, whereas wind speed proved not be significantly associated (Oliveiros et al. 2020). Based on Shi et al. conclusions, lower and higher temperature rates may decrease the COVID-19 incidence rates and the role of absolute humidity has not yet been established (Shi et al. 2020). Lau et al. recorded an increase in the doubling time of COVID-cases and this was attributed to the lockdown measurements implemented (Lau et al. 2020). Chen et al. found out that the optimal temperature for COVID-19 is 8.07 °C, within a humidity range of 60-90% (Chen et al. 2020). Sun et al. concluded that cold and dry winter are considered as a common environmental condition conductive for COVID-19 (Sun et al. 2020). Peeri et al. attributed the increased and rapid COVID-19 perforation to air travel frequency and circumstances (i.e. connection flights) (Peeri et al. 2020). Gostic et al. estimated that screening during travel may miss more than half of the infected cases, as the have not developed symptoms in the time of screening (Gostic et al. 2020). Sajadi et al. reached the conclusion that temperature range of 5-11°C, combined with low specific range of 3-6g/kg and absolute humidity range of 4-7 g/kg are the optimal environmental factors for COVID-19 transmission (Sajadi, Habibzadeh, Vintzileos, Shokouhi, et al. 2020). Qu et al. linked the COVID-19 transmission with airborne dust (Qu et al. 2020). Gupta showed that for every 1°C increase above 5 °C, the temperature as factor may decrease the COVID-19 transmission rate by 10% (Gupta 2020). Cai et al. found no correlation between the daily mean temperature and the epidemic growth rate in case of Hunan or Wuhan, but insist that there is a weak correlation between the daily mean temperature and the mortality rates in both provinces (Cai et al. 2020). Chinazzi et al. assessed travel limitations practices applied in China and international scale and verified travel quarantine delayed the epidemic progression 3 to 5 days in China or more in worldwide basis (Chinazzi et al. 2020). Lee et al. reviewed Singapore’s approach to COVID-19 epidemic concerning travel restrictions applied at all ports of entry (Lee, Chiew, and Khong 2020). Wang et al. propose that high temperature and high relative humidity significantly affects the COVID-19 transmission rates (J. Wang et al. 2020). Poole suggests that a climatological range of 4-12°C within an area of 25-55° latitude may enhance the COVID-19 spread (Poole 2020). Bu et al. conclude that temperature rate of 13-19°C and humidity rate of 50-80% are conducive to the virus survival (Bu et al. 2020).

### Study quality

The studies included in this systematic review were scored from 17 to 19.8, upon the criteria predefined. The criteria on which studies were assessed with the minimum score were related to their not clearly addressing the following items: report of the study design and assessing method in the title and abstract; clearly define the participants, the interventions and the outcomes; clearly state handle of missing data and accuracy of data; generalization of the findings.

## Discussion

To the best of our knowledge, the present systematic review is the first to summarize the available evidence on the association of COVID-19 with environmental factors. Taking into consideration that the new coronavirus is a new human pathogen, which due to its outbreak in China and its rapid spread worldwide, it is important to understand reliable epidemiological information for its survival in the environment (Chen et al. 2020). Therefore, it is necessary to find prognostic predictors to distinguish high-risk areas or countries and improve the new challenging situation.

Any infectious disease origins and spread occur only when affected by certain natural and social factors through acting on the source of infection, the mode of transmission and the susceptibility of the population. The environmental factors such as meteorological factors, namely temperature and humidity proved to play a part in coronavirus outbreak besides the social factors (Jia et al. 2020). In our systematic review, overall evidence is sufficiently robust to determine the impact of temperature in virus’ survival via different methods, like the effect of each 1°C increase which lowers the virus’ R by 0.225 (J. Wang et al. 2020) and the doubling time of the confirmed cases which is positively correlated with temperature (Oliveiros et al. 2020). Four, studies included in this systematic review determine the exact temperature range, within which temperature is conducive to virus spread and survival. The pooled results of these 4 studies indicated that temperature range different from 4-24°C is not conducive to the survival of the coronavirus (M. Wang et al. 2020) (Sajadi, Habibzadeh, Vintzileos, Miralles-wilhelm, et al. 2020) (Poole 2020) (Bu et al. 2020). Concerning humidity, although the results in this review did not reveal robust associations between humidity and coronavirus survival and are always validated in combination with temperature, they need to be interpreted carefully given the monotonic functional relationship between humidity and temperature. In other words, if temperature was associated to COVID-19 transmission, very likely absolute humidity would play a role. Pooled results of the studies included in this systematic review show that combined with high temperature, absolute humidity range of 4-7 g/m^3^ (Sajadi, Habibzadeh, Vintzileos, Miralles-wilhelm, et al. 2020) or specific humidity range of 3-6 g/kg (Sajadi, Habibzadeh, Vintzileos, Miralles-wilhelm, et al. 2020) or humidity of 50-80% (Sajadi, Habibzadeh, Vintzileos, Miralles-wilhelm, et al. 2020) may reduce the transmission of COVID-19. Other factors concerned, such as air index, rainfall/precipitation, wind speed, do not show to have significant impact to virus stability and survival and need to be further assessed. Although not all environmental factors and not in depth are clearly described by authors of the included studies, important associations are observed and need further investigation. Variability in the results among the studies included in our review may be attributed to i) the utilization of different types of assessing methods of each environmental factor, ii) the different qualitative characteristics of the populations used, iii) sample size, iv) duration of the study and others.

Environmental factors, characterized by lag effects and threshold effects, can target at two objects, host and virus, during infectious disease outbreak. On one hand, human activity patterns and immunity can be influenced by environmental factors. But the effect caused by environmental condition was limited during the COVID-19 outbreak, due to the absence of extreme weather and specific immunity for a newly emerging virus. On the other hand, environmental impacts on the SARS-CoV-2 are more significant than the host population because the transmission and virulence of the virus varies in different conditions. Finally, environmental impacts on transmission of virus should be characterized in the dynamic model, because infectiousness estimated in the traditional dynamic model is actually a confounding effect with environmental effect. It is necessary to take account of environmental issues based on dynamic transmission model so that the impacts could be isolated and qualified.

Apart from the he basic strengths of this review regards the study quality assessment, which was based on STROBE, its PRISMA compliance approach and the peer-review process followed.

## Limitations

Due to limited data available, other meteorological factors such as air pressure, atmospheric particles, ultraviolet, and social factors such as population movement were not included for analysis. Inclusion of such factors will provide more accurate and reliable results.

In addition, the relatively short time length of the current outbreak, combined with imperfect daily reporting practices, make our results vulnerable to changes as more data becomes available. We have assumed that travel limitations and other containment interventions have been implemented consistently across provinces and have had similar impacts (thus population mixing and contact rates are assumed to be comparable), and have ignored the fact that different places may have different reporting practices. Further improvements could incorporate data augmentation techniques that may be able to produce historical time series with likely estimates of case counts based on onset of disease rather than reporting dates. This, along with more detailed estimates of the serial interval distribution, could yield more realistic estimates of R. Finally, further experimental work needs to be conducted to better understand the mechanisms of transmission of COVID-19. Mechanistic understanding of transmission could lead to a coherent justification of our findings.

## Conclusions

In summary, this review provided evidence that high temperature and high humidity reduce the COVID-19 transmission. However, further studies concerning other environmental (namely meteorological) factors’ role should be conducted in order to further prove this correlation.

## Data Availability

The data is available if needed

